# Genome-wide association study of susceptibility to pneumococcal carriage amongst children

**DOI:** 10.64898/2026.07.13.26356474

**Authors:** Rama Kandasamy, Meeru Gurung, Sonu Shrestha, Sagida Bibi, Stephen Thorson, Michael Carter, Daniel O’Connor, David R. Murdoch, Dominic F. Kelly, Shrijana Shrestha, Michael Levin, Andrew J. Pollard

**Affiliations:** Discipline of Child and Adolescent Health, Faculty of Medicine and Health, The University of Sydney, Australia; National Centre for Immunisation Research and Surveillance, The Children’s Hospital at Westmead, Australia; Paediatric Research Unit, Patan Academy of Health Sciences, Kathmandu, Nepal; Oxford Vaccine Group, Department of Paediatrics, University of Oxford, Oxford, United Kingdom; NIHR Oxford Biomedical Research Centre, Oxford, United Kingdom; Centre for Human Genetics, University of Oxford, Oxford, United Kingdom; Paediatric Intensive Care Unit, Oxford University Hospitals NHS Foundation Trust, Oxford, United Kingdom; Department of Pathology, University of Otago, Christchurch, New Zealand; Department of Infectious Disease, Faculty of Medicine, Imperial College London, United Kingdom

**Keywords:** Nepal, children, carriage, pneumococcus, GWAS

## Abstract

**Background:** Pneumococcal disease is a leading cause of paediatric pneumonia and meningitis. Pneumococcal colonisation is the fundamental step to pneumococcal disease causation. We aimed to identify genetic loci associated with pneumococcal colonisation amongst children.

**Methods:** We conducted a genome-wide association study on 2111 Nepalese children, comprising 1346 cases carrying pneumococcus and 765 controls. We tested 8.1 million imputed variants using logistic regression and ten principal components as covariates. Fine mapping and functional evidence were used to identify suspected causal variants and related genes of interest.

**Findings:** A cluster of 22 variants of genome-wide significance (p<5×10^-8^) were identified on chromosome 12q21.31, eight of which were within *PPFIA2*. Fine mapping of this region identified 5 variants within 0.1 Mb of the 5’ region of *PPFIA2* all of which are significant eQTLs for PPFIA2. We further describe three loci (10q23.31, 12q23.1, and 20p11.21) which had variants with highly suggestive associations (p<5×10^-7^)with pneumococcal carriage.

**Interpretation:** Our study demonstrate human susceptibility to pneumococcal carriage to be polygenic with genetic variations which regulate *PPFIA2* expression playing a key role in the ability for pneumococcus to colonise children. Targeting these genetic factors and the associated pathways are a means for preventing pneumococcal disease.

**Funding:** This study was supported by funding from Gavi - the vaccine alliance, the European Union’s Horizon 2020 research and innovation program under grant agreement number 668303 (PERFORM), and a Robert Austrian Research Award.

**Research in context:** *Evidence before this study:* Pneumococcus, through the causation of pneumonia and invasive disease, is a common infectious cause of death in children. Pneumococcal colonisation is the seminal step to disease development. Yet, there are limited data on how human genetics influence pneumococcal carriage.

*Added value of this study:* The study examined the genetic association of pneumococcal carriage in 1346 healthy children and 765 controls with four genetic regions of interest prioritised. Fine mapping and functional analyses of these regions support *PPFIA2, HRT7, NTN4*, and *FOXA2* as genes of interest for susceptibility to pneumococcal colonisation.

*Implications of all the available evidence:* Our study shows a link between multiple genes and pneumococcal colonisation. Improved understanding of the action of these genes in the respiratory tract may inform novel approaches to disease prevention.

## INTRODUCTION

Pneumococcus was responsible for an estimated 154,773 deaths in children under 5 years of age across the world in 2021.(1) Pneumococcus frequently asymptomatically colonises the upper respiratory tract (carriage) of children and in a small number of cases migrates to the lower respiratory tract to cause pneumonia or into normally sterile sites such as the blood stream and the meninges to cause sepsis and meningitis (invasive pneumococcal disease).(2) Vaccination against pneumococcal disease using pneumococcal conjugate vaccines (PCVs) has broadly been successful however, there are limitations. Notably many settings have observed an increase in pneumonia and invasive pneumococcal disease (IPD) which is related to pneumococcal serotypes not covered by the current PCVs, diminishing the nett benefits of the vaccines.(3) Whilst PCVs which have been designed to cover more serotypes have been found to be less immunogenic and potentially have reduced protection.(4) As such there is a pressing need to develop new preventative strategies. Given that carriage is the fundamental step to disease causation, better understanding of the host factors responsible for colonisation is key to progressing novel disease prevention approaches.

Human genome-wide association studies (GWAS) are a powerful approach to better understanding susceptibility to infectious diseases. This is evidenced by a growing body of studies which describe different human genetic factors playing a role in susceptibility, severity and outcome of specific infections.(5) Such information can be used to identify those who are at increased risk and inform the development of new interventions. This approach is exemplified by a GWAS of invasive meningococcal disease which showed that variants within *CFHR3 (complement factor H related 3)* are associated with disease.(6) Subsequent functional studies showed that these *CFHR3* variants alter the concentration of complement factor H which in turn alters a person’s ability to kill meningococcus.(7) These findings supported the selection of the meningococcal protein FHBP (factor H binding protein, which binds human CFHR3), as a vaccine antigen. Two vaccines containing this antigen are now licensed and are the only available vaccines against meningococcal group B disease.(8, 9) Additionally, the findings have supported additional clinical decision making, with people who have complement disorders or who are on medications which effect complement function, now being recommended to receive additional doses of meningococcal vaccines.(10)

We have previously conducted a pneumococcal carriage study amongst Nepalese children.(11) These children underwent nasopharyngeal sampling and analysis for the presence of pneumococcus. As part of this study, we also collected samples for human DNA extraction and genotyping. We aimed to conduct a GWAS in this cohort to determine whether there are host genetic factors associated with pneumococcal carriage.

## METHODS

### Study design

We conducted a GWAS of healthy community-based Nepalese children who were assigned to case and control cohorts based on the detection of pneumococcus on a nasopharyngeal swab. The study was approved by the Nepal Health research Council (Reg. no. 286/2014) and Oxford Tropical Research Ethics Committee (OxTREC Reference: 11-15).

### Sample processing

Nasopharyngeal swabs were collected, cultured for pneumococcus, and serotyped as previously described.(11) DNA was extracted from saliva samples collected from participating children using the Oragene DNA Sample Collection Kit, OG-250 Disc Format (Genotek, Ottawa, Ontario, Canada). Saliva samples were incubated at 50-60 °C prior to having DNA extracted using the QIAsymphony platform using QIAsymphony DSP DNA midi kits.

### Genotyping analysis

DNA was genotyped using the Illumina Global Screening array. Filtering criteria of the generated data included sample and SNP call rates > 95%, minor allele frequency < 5%, Hardy-Weinberg equilibrium with p < 10^-5^, elevated missingness (> 0.2 for individuals and variants), heterozygosity outliers, relatedness, and discrepancies between genetically determined and reported sex. The genetic data were merged with the 1000 Genomes data to screen for ethnic outliers using the top two principal components.

### Imputation

Genetic data were imputed on the Michigan Imputation Server using the 1000 Genomes phase 3 v5 reference panel, and Eagle v2.4 phasing. Imputed data then underwent a further quality control step where SNPS of low imputation quality (r-squared < 0.3), triallelic SNPs, and minor allele frequency < 1% were removed.

### Colocalisations

The coloc package, coloc.abf function, was used to estimate colocalisations between GTEx V10 eQTLs data across all tissues and the sentinel variants using a 500 kb window. Colocalisations with a posterior probability, PP.H4, >0.8 were considered significant.

### Fine mapping

Fine mapping of regions of interest was conducted using “Sum of Single Effects” (SuSiE) and the corresponding LD matrix. 95% credible sets of variants were identified and posterior inclusion probabilities (PIP) calculated to determine the probability for a SNP to be one of the causal SNPs in the region.

### Functional genomic analyses

The functional impact of variants of interest was explored using the variant annotation databases; RegulomeDB V2.2, Combined Annotation Dependent Depletion (CADD) v1.7 (CADD model GRCh38-v1.7), and HaploReg v4.2.(12-14) Significant eQTLs were determined for variants of interest using GTEx V10, with multiple significant values at the same site across multiple tissues collapsed to the smallest value for visualisation purposes.(15) Single cell gene expression data were explored for the genes of interest using CZ CellxGene Discover, https://cellxgene.cziscience.com/gene-expression, accessed on 9^th^ March 2026, after filtering for normal, human, lung tissue samples.(16) 36 data sets remained after initial filtering with these gene expression data then filtered for cell counts greater than 1000 and the top ten cell types by gene expression level (normalised read counts) for each gene of interest.

### Statistical analyses

Genetic data preparation, filtering, and analysis was conducted using PLINK 2.0. Data underwent pre-imputation quality checks and preparation using McCarthy Group Tools.(17) A GWAS on 8,112,140 variants, using the presence of pneumococcal carriage as a binary phenotype (1346 cases and 765 controls) was conducted using logistic regression with the top ten principal components as covariates in PLINK 2.0.(18) Summary statistics were upload to LocusZoom to generate locus specific visualisations. All other analyses and visualisations were conducted in RStudio 2024.12.1.

## RESULTS

### GWAS findings

A genome-wide association study of pneumococcal carriage amongst Nepalese children with 1346 cases and 765 controls was conducted across a total of 8,112,140 variants (figure 1). The demographic characteristics of the study participants indicated sex distribution and median ages between the groups were comparable (table 1). The most frequently carried pneumococcal serotypes amongst the carriage group were non-typeable, serotype 6A, and serotype 6B (supplementary table 1).

**Table 1.** Participant characteristics.

|  | **Controls N=765** | **Cases N=1346** |
| --- | --- | --- |
| Male, N (%) | 423 (55.3) | 711 (52.3) |
| Age, median years (IQR) | 1.1 (0.8-1.4) | 1.1 (0.8-1.5) |

**Table 2.** Sentinel variants associated with pneumococcal carriage.

| SNP | Chromosome | Position | Ref | ALT | OR (95% CI) | p-value | MAF | Nearest gene |
| --- | --- | --- | --- | --- | --- | --- | --- | --- |
| rs9634233 | 12 | 81861185 | G | A | 0.51 (0.41-0.64) | $2.38 \times 10^{-9}$ | 0.12 | PPFIA2 |
| rs17098642 | 10 | 90681086 | A | G | 0.62 (0.52-0.75) | $2.74 \times 10^{-7}$ | 0.19 | HTR7 |
| rs75742107 | 12 | 95766618 | G | A | 0.56 (0.44-0.71) | $1.33 \times 10^{-6}$ | 0.08 | NTN4 |
| rs1203884 | 20 | 22563062 | A | G | 0.64 (0.1-0.53) | $2.19 \times 10^{-6}$ | 0.14 | FOXA2 |

**Figure 1.**
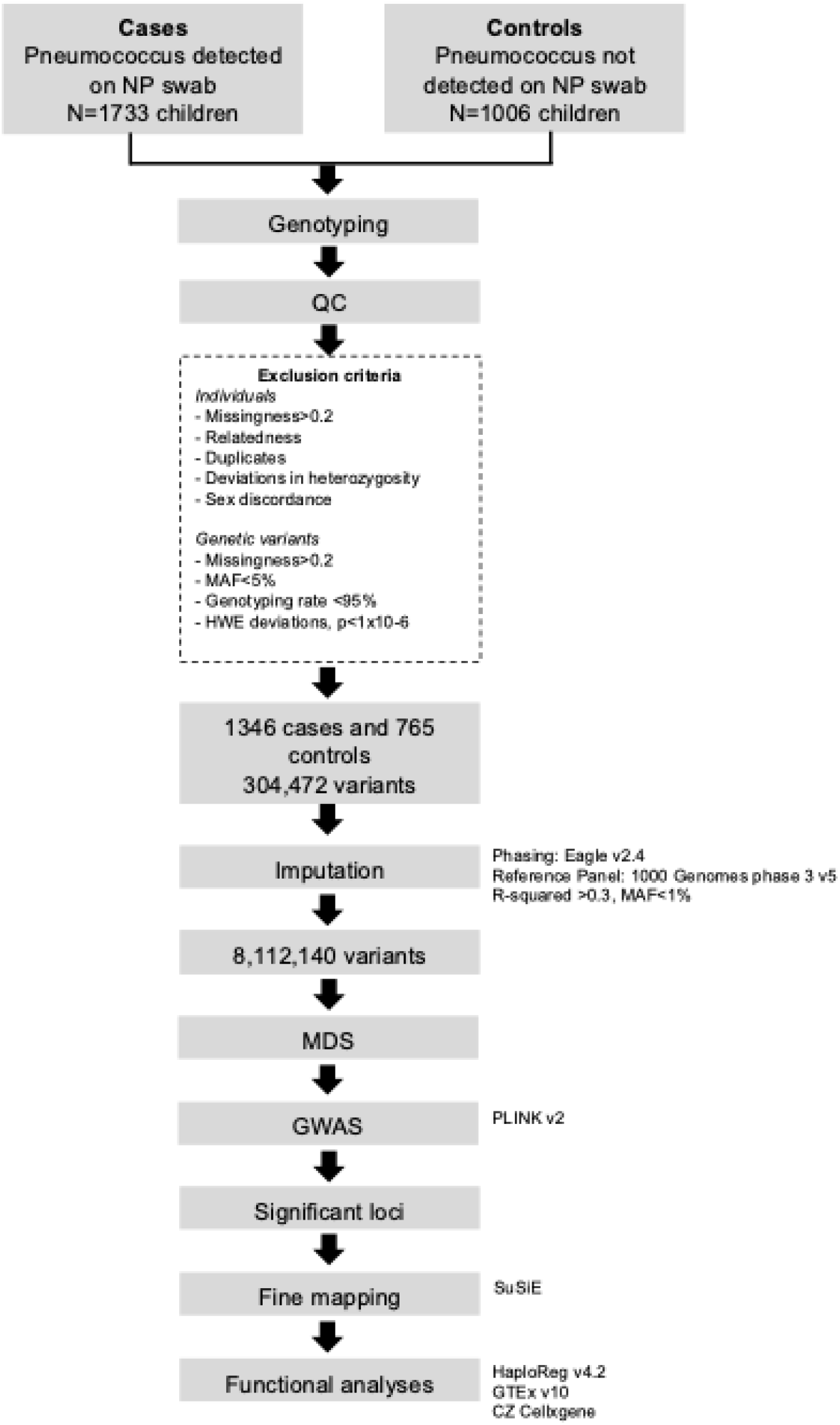
GWAS design flow chart.

A lack of genomic inflation was demonstrated by a 50^th^ centile lambda of 1.008 (supplementary figure 1). Association analysis revealed 22 genome-wide significant variants which were located in a single region on chromosome 12 (figure 2A, supplementary table 2), with 8 of these variants intronic to *PPFIA2* and the remainder intergenic to *PPFIA2* and *CCDC59* (figure 2B). Variants on chromosomes 10q23.31, 12q23.1, and 20p11.21, related to the genes *HRT7, NTN4*, and *FOXA2* respectively were highly suggestive of an association with pneumococcal carriage.

**Figure 2.**
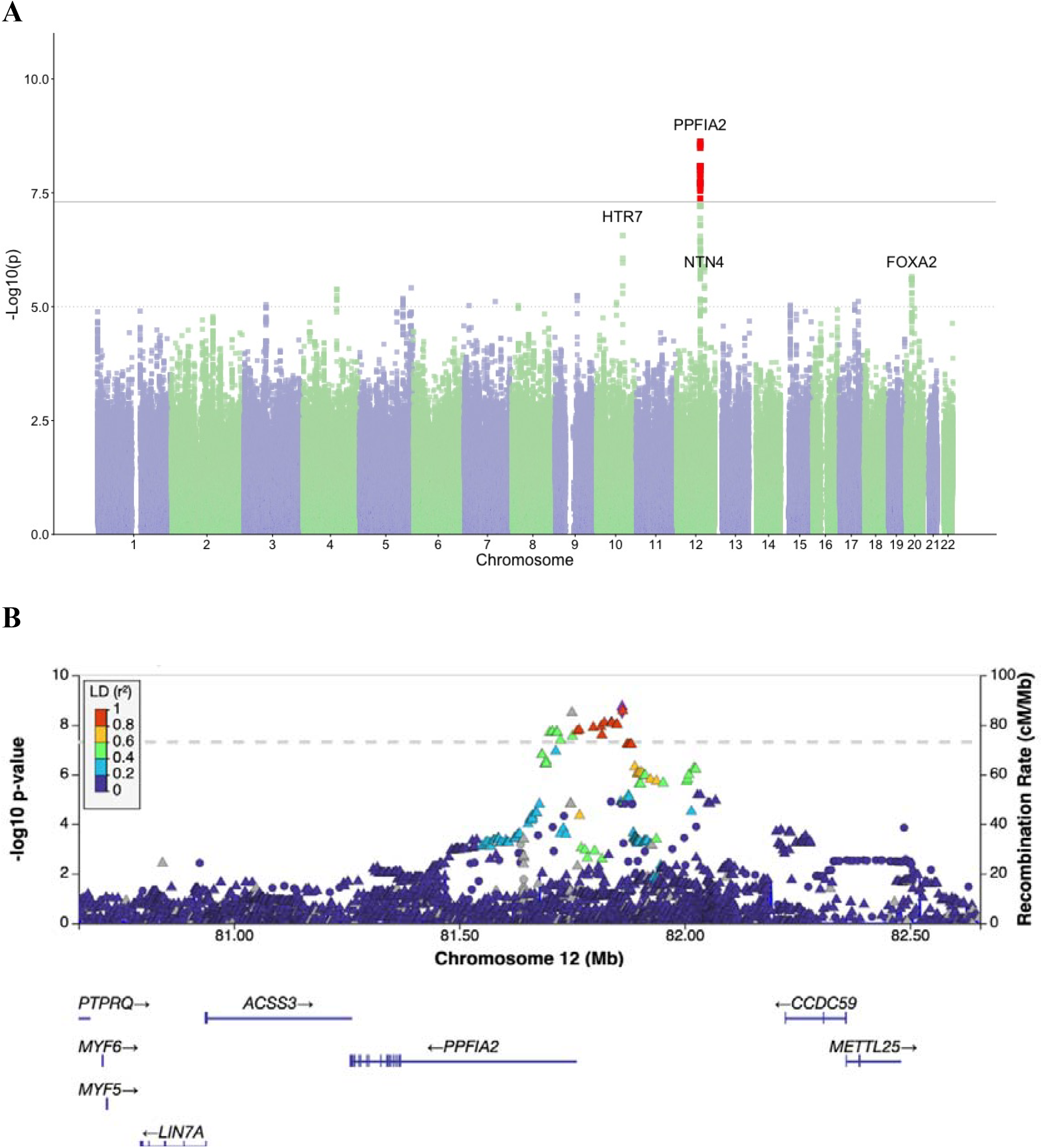
Manhattan plot (A) and locus zoom plot (B) of genome-wide significant variant peak on chromosome 12.

### Colocalisation

No significant colocalisations between the lead variant at the *PPFIA2* locus (12q21.31) and gene expression in the analysed tissues were identified. Notably there was a high probability that both GWAS and eQTL traits are associated, but with different causal variants (i.e. PP.H3>0.8) for PPFIA2. No significant colocalisations were identified for the lead variants at 10q23.31, 12q23.1, and 20p11.21. At both 10q23.31 and 20p11.21 there were high probabilities that both GWAS and eQTL traits are associated, but with different causal variants (supplementary figures 2-5).

### Fine-mapping

Fine mapping of the lead variant locus on chromosome 12 was able to delineate two credible sets of three and two variants each (table 3). All credible set variants were intergenic and within 0.1 Mb of the 5’ region of *PPFIA2*. The variant rs4842301 had the highest posterior inclusion probability. Fine mapping of the *HTR7* locus on chromosome 10 identified 9 variants across 7 credible sets (supplementary table 3). Six of these variants had a posterior inclusion probability of 1. All credible set variants were intergenic and within 0.1 Mb of the 3’ region of *HTR7*. Fine mapping of the *NTN4* locus on chromosome 12 identified 9 variants across 9 credible sets, with 8/9 variants intronic to *NTN4* (supplementary table 4). Fine mapping of the *FOXA2* locus on chromosome 20 identified 5 variants across 5 credible sets. All variants were intergenic and within 0.1 Mb of the 3’ region *FOXA2* (supplementary table 5).

**Table 3.** Credible sets from fine mapping of the variants proximal to PPFIA2 and the associated functional annotations.

| rsID | MAF | p-value | OR (95% CI) | Credible set | PIP | RegulomeDB score | RegulomeDB rank | CADD score |
| --- | --- | --- | --- | --- | --- | --- | --- | --- |
| rs9634233 | 0.12 | 2.38x10<sup>-9</sup> | 0.51 (0.41-0.64) | Sentinel variant | - | 0.60906 | 4 | 1.127 |
| rs2471500 | 0.09 | 8.17x10<sup>-9</sup> | 0.51 (0.41-0.64) | 1 | 0.9175409 | 0 | 5 | 3.92 |
| rs2464738 | 0.09 | 8.17x10<sup>-9</sup> | 0.51 (0.41-0.64) | 1 | 0.9175409 | 0.27598 | 6 | 1.92 |
| rs7134579 | 0.09 | 8.19x10<sup>-9</sup> | 0.51 (0.41-0.64) | 1 | 0.6964975 | 0.14536 | 6 | 8 |
| rs4842301 | 0.12 | 9.80x10<sup>-9</sup> | 0.51 (0.41-0.65) | 2 | 0.9998311 | 0.18412 | 7 | 5.069 |
| rs10083046 | 0.12 | 9.88x10<sup>-9</sup> | 0.51 (0.41-0.65) | 2 | 0.6201584 | 0.18412 | 7 | 1.853 |

PIP=posterior inclusion probability. CADD=Combined Annotation Dependent Depletion (higher score represent more likely to be deleterious)

### Functional analyses

At the *PPFIA2* locus rs4842301 had the highest Combined Annotation Dependent Depletion (CADD) score. All of the credible set variants from the *PPFIA2* locus demonstrated significant eQTL associations with *PPFIA2* across five different tissue types; oesophageal muscle, mammary tissue, adipose tissue, tibial nerve, and cultured fibroblasts (supplementary figure 6A and supplementary table 6). PPFIA2 expression amongst respiratory tract tissues was found to be highest in helper T cells, alveolar capillary type 2 endothelial cells, and efferent neurons (suppplementary 6B). All of the credible set variants from the *PPFIA2* locus were reported to alter regulatory motifs (supplementary table 7).

At the *HTR7* locus rs2710725 had the highest CADD score. Of the credible set variants from the *HTR7* locus 7/8 demonstrated a total of 28 significant eQTL associations. The majority, 24/28, of these significant eQTLs were to *HTR7* and spanned across seven different tissue types (supplementary figure 7A). HTR7 expression amongst respiratory tract tissues was highest amongst stromal cells, lung ciliated cells, and secondary crest myofibroblasts (supplementary figure 7B).

At the *NTN4* locus the sentinel variant rs75742107 had the highest CADD score followed by rs78879109. There were four credible set variants which had 78 significant eQTLs identified across seven genes, *AMDHD1* (30/78), *SNRPF* (18/78), *SNRPF-DT* (10/78), *ENSG00000257878* (7/78), *NTN4* (5/78), *CCDC38* (4/78), and *HAL* (4/78). Significant eQTLs for *AMDHD1* were identified across eleven different tissue types (supplementary figure 8A). AMDHD1 expression amongst respiratory tract tissues was highest amongst neutrophils and CD4-positive T cells (supplementary figure 8B).

At the *FOXA2* locus the sentinel variant rs1203884 had the highest CADD score followed by rs6137690. A single credible set variant, rs6137690, had a single significant eQTL for the uncharacterised gene *ENSG00000283072* amongst tibial artery tissue. FOXA2 expression amongst respiratory tract tissues was highest amongst mesenchymal stem cells and mesothelial cells (supplementary figure 9).

## DISCUSSION

We report the first GWAS of pneumococcal carriage and have identified 22 variants on chromosome 12 (12q21.31) which reach genome-wide significance. Functional analyses indicate that variants at this locus regulate the expression of PPFIA2. We further describe variants at three loci (10q23.31, 12q23.1, 20p11.21), related to *HTR7, NTN4*, and *FOXA2*, which have a suggestive association with pneumococcal carriage.

PPFIA2 has a well described function as a pre-synaptic scaffolding protein facilitating synaptic vesicle release in neuronal cells within the central nervous system.(19) However, the function of PPFIA2 peripherally in the respiratory tract has not been described. Genetic variants of *PPFIA2* have previously been found to be associated with a broad range of traits, most notably of which in the context of this study are changes in lung function, gut microbiome variations, and COVID-19 severity.(20) Interestingly, *PPFIA2* expression has been associated with SARS-CoV-2 viral load in the upper respiratory tract and susceptibility to asthma exacerbations.(21, 22) Given the well described function of PPFIA2 in the nervous system and the high expression levels described in efferent neuron cells of the respiratory tract it is possible that PPFIA2 modifies the ability of pneumococcus to colonise through neurologically mediated processes in the respiratory tract. Further investigation into the effect PPFIA2 may have on nervous system regulation in the respiratory tract is required.

The serotonin receptor 5-HT_7_ is encoded by *HTR7*, with 5-HT_7_ shown to regulate inflammatory responses on human airway epithelial cells.(23) Interestingly ciliated cells were one of the respiratory tract cell lines shown to have high HTR7 expression with a prior study of mice showing that serotonin increases cilia driven particle transport in the trachea.(24) Serotonin receptors have also been shown to be involved in viral entry to host cells, for a broad range of different viruses, with a drug designed to inhibit the 5-HT_7_ receptor shown to inhibit the endocytosis of influenza virus *in vitro*.(25) Additionally, there are some data to indicate that serotonin can directly act upon bacteria stimulating quorum sensing and enhanced virulence.

*NTN4* encodes a protein from the netrin family which has been reported to have a role in axon guidance, angiogenesis, and lung morphogenesis.(26) Genetic variants of NTN4 have been implicated in COVID-19 severity and lung function.(20) We note that in this present study the credible set variants identified had a number of eQTLs with *AMDHD1* rather than *NTN4*. AMDHD1 has been reported to be a key enzyme in histidine degradation whilst genetic variants have been associated with vitamin D levels. Interestingly there are a growing body of data linking vitamin D levels to susceptibility to infectious diseases.

*FOXA2* encodes a forkhead class of transcription factor which has a role in lung development and mucous homeostasis. FOXA2 conducts mucous homeostasis through the regulation of the mucin genes *MUC5AC* and *MUC5B*.(27) Airway pathogens such as *P. aeruginosa* have been shown to express virulence factors which inhibit FOXA2 activity leading to disruption of mucous homeostasis and bacterial colonisation.(28) Interestingly variants proximal to *MUC5AC* have been reported as having a genome-wide significant association with susceptibility to pneumonia.(29) Further exploration into whether pneumococcus possesses virulence factors which modify FOXA2 expression and mucous homeostasis are required.

Limitations of this study include the absence of comparable analyses in non-Nepalese populations and the potential influence of unascertained biases (e.g. smoke exposure). Further replication of our findings in those of non-Nepalese ancestry would allow greater confidence in generalising them to all children.

In conclusion we have shown that susceptibility to pneumococcal carriage is heritable and polygenic. Further replication of these findings in alternate ethnic backgrounds and exploration of the effects variations of these loci have on respiratory tract function are required.

## Data Availability

All data produced in the present study are available upon reasonable request to the authors

