## supplement for "Genome-wide association study of susceptibility to pneumococcal carriage amongst children"

**Supplementary information**

**Supplementary Table 1. Pneumococcal serotypes carried by Nepalese children included in the genome-wide association study.**

| **Serotype** | **Number** |
| --- | --- |
| NT | 147 |
| 6A | 69 |
| 6B | 66 |
| 11A | 60 |
| 19F | 58 |
| 15B | 53 |
| 34 | 53 |
| 14 | 50 |
| 6C | 50 |
| 23F | 48 |
| 10A | 47 |
| 19A | 47 |
| 15C | 44 |
| 13 | 36 |
| 33B | 30 |
| 35B | 30 |
| 9V | 28 |
| 16F | 25 |
| 15A | 24 |
| 23A | 24 |
| 35A | 23 |
| 23B | 22 |
| 35F | 22 |
| 21 | 21 |
| 17F | 15 |
| 18C | 15 |
| 19B | 14 |
| 20 | 13 |
| 31 | 13 |
| 24F | 12 |
| 33F | 10 |
| 4 | 10 |
| 8 | 10 |
| 38 | 9 |
| 6D | 9 |
| 7B | 9 |
| 10F | 8 |
| 18A | 8 |
| 22F | 8 |
| 28F | 8 |
| 35C | 8 |
| 1 | 6 |
| 39 | 6 |
| 7C | 6 |
| 18F | 5 |
| 42 | 5 |
| 5 | 5 |
| 7F | 5 |
| 10B | 4 |
| 15F | 4 |
| 24A | 4 |
| 11B | 3 |
| 29 | 3 |
| 3 | 3 |
| 33C | 3 |
| 33D | 3 |
| 48 | 3 |
| 9N | 3 |
| 12F | 2 |
| 17A | 2 |
| 22A | 2 |
| 25A | 2 |
| 33 | 2 |
| 16B | 1 |
| 19C | 1 |
| 28A | 1 |
| 32F | 1 |
| 36 | 1 |
| 37 | 1 |
| 40 | 1 |
| 7A | 1 |
| 9L | 1 |

**Supplementary Table 2. Variants of genome wide significance identified in a GWAS of pneumococcal carriage**

| **rsID** | **MAF** | **p-value** | **OR (95% CI)** |
| --- | --- | --- | --- |
| rs73149327 | 0.060815 | 2.00x10^-08^ | 0.40 (0.29-0.55) |
| rs138006051 | 0.0605346 | 2.03x10^-08^ | 0.40 (0.29-0.55) |
| rs377625270 | 0.0616979 | 1.86x10^-08^ | 0.40 (0.29-0.55) |
| rs73151310 | 0.0622474 | 1.88x10^-08^ | 0.40 (0.29-0.55) |
| rs73151314 | 0.0613134 | 2.02x10^-08^ | 0.40 (0.29-0.55) |
| rs12426253 | 0.0584164 | 4.19x10^-08^ | 0.41 (0.29-0.56) |
| rs2464760 | 0.0759288 | 3.24x10^-09^ | 0.41 (0.30-0.55) |
| rs2464759 | 0.0612012 | 2.84x10^-08^ | 0.42 (0.30-0.57) |
| rs2453072 | 0.100947 | 1.82x10^-08^ | 0.52 (0.41-0.65) |
| rs2464750 | 0.100206 | 1.62x10^-08^ | 0.52 (0.42-0.65) |
| rs2464748 | 0.0996669 | 1.27x10^-08^ | 0.52 (0.41-0.65) |
| rs2464745 | 0.0994619 | 1.19x10^-08^ | 0.52 (0.41-0.65) |
| rs2471497 | 0.100616 | 2.54x10^-08^ | 0.53 (0.42-0.66) |
| rs7134579 | 0.0980887 | 8.19x10^-09^ | 0.51 (0.41-0.64) |
| rs2471501 | 0.0979513 | 8.41x10^-09^ | 0.51 (0.41-0.64) |
| rs2471500 | 0.0980846 | 8.17x10^-09^ | 0.51 (0.41-0.64) |
| rs2464738 | 0.0980846 | 8.17x10^-09^ | 0.51 (0.41-0.64) |
| rs2471499 | 0.0988187 | 8.47x10^-09^ | 0.51 (0.41-0.65) |
| rs10083046 | 0.118679 | 9.88x10^-09^ | 0.51 (0.41-0.65) |
| rs4842301 | 0.118914 | 9.80x10^-09^ | 0.51 (0.41-0.65) |
| rs9634233 | 0.122611 | 2.38x10^-09^ | 0.51 (0.41-0.64) |
| rs6539586 | 0.122784 | 2.77x10^-09^ | 0.51 (0.41-0.64) |

**Supplementary Table 3. Credible sets from fine mapping of the variants proximal to HTR7 and the associated functional annotations.**

| **rsID** | **MAF** | **p-value** | **OR (CI 95%)** | **Credible set** | **PIP** | **RegulomeDB score** | **RegulomeDB rank** | **CADD score** |
| --- | --- | --- | --- | --- | --- | --- | --- | --- |
| rs17098642 | 0.18 | 2.74x10^-7^ | 0.69 (0.6-0.8) | Sentinel variant | - | 0.18412 | 7 | 0.466 |
| rs2710717 | 0.4 | 4.01x10^-2^ | 0.85 (0.73-0.99) | 7 | 0.3730255 | 0.51392 | 7 | 5.192 |
| rs2710725 | 0.4 | 4.01x10^-2^ | 0.85 (0.73-0.99) | 7 | 0.3134873 | 0.55324 | 1f | 5.422 |
| rs2710726 | 0.4 | 4.01x10^-2^ | 0.85 (0.73-0.99) | 7 | 0.3134873 | 0.55324 | 1f | 4.655 |
| rs12246059 | 0.19 | 4.16x10^-2^ | 1.2 (1.01-1.43) | 9 | 1 | 0.22271 | 1f | 0.112 |
| rs1419351 | 0.32 | 3.5x10^-6^ | 0.72 (0.63-0.83) | 1 | 1 | 0.1574 | 6 | 1.462 |
| rs1419350 | 0.3 | 3.52x10^-2^ | 1.18 (1.01-1.37) | 8 | 1 | 0.22267 | 6 | 5.068 |
| rs2901088 | 0.3 | 8.65x10^-7^ | 0.69 (0.6-0.8) | 6 | 1 | 0.51392 | 7 | 0.015 |
| rs1892107 | 0.43 | 2.47x10^-5^ | 0.74 (0.64-0.85) | 2 | 1 | 0.51392 | 7 | 5.194 |
| rs1892105 | 0.13 | 4.74x10^-1^ | 1.08 (0.87-1.35) | 4 | 1 | 0.51392 | 7 | 4.732 |

**Supplementary Table 4. Credible sets from fine mapping of the variants proximal to NTN4 and the associated functional annotations.**

| **rsID** | **MAF** | **p-value** | **OR (CI 95%)** | **Credible set** | **PIP** | **RegulomeDB score** | **RegulomeDB rank** | **CADD score** |
| --- | --- | --- | --- | --- | --- | --- | --- | --- |
| rs75742107 | 0.08 | 1.33x10^-6^ | 0.56 (0.44-0.71) | Sentinel variant | - | 0.60906 | 4 | 10.87 |
| rs11108232 | 0.23 | 4.06x10^-1^ | 0.93 (0.8- 1.1) | 8 | 1 | 0.51392 | 7 | 0.584 |
| rs191318919 | 0.07 | 6.52x10^-5^ | 0.58 (0.44-0.76) | 10 | 1 | 0.18412 | 7 | 0.737 |
| rs35202071 | 0.14 | 2.25x10^-1^ | 1.13 (0.93-1.38) | 7 | 1 | 0.51392 | 7 | 1.916 |
| rs55784519 | 0.35 | 3.45x10^-2^ | 1.17(1.01-1.35) | 6 | 1 | 0.18412 | 7 | 1.916 |
| rs78879109 | 0.09 | 7.22x10^-6^ | 0.60 (0.48-0.75) | 4 | 1 | 0.18412 | 7 | 7.398 |
| rs139424288 | 0.09 | 7.15x10^-6^ | 0.60 (0.48-0.75) | 1 | 1 | 0.60906 | 4 | 1.273 |
| rs7303360 | 0.28 | 9.87x10^-1^ | 1 (0.86-1.17) | 3 | 1 | 0.55324 | 1f | 0.279 |
| rs10859948 | 0.37 | 4.49x10^-3^ | 0.81 (0.71-0.94) | 2 | 1 | 0.55436 | 1f | 0.814 |
| rs151258214 | 0.03 | 1.23x10^-2^ | 0.61 (0.41-0.9) | 9 | 1 | 0.60906 | 4 | 2.431 |

**Supplementary Table 5. Credible sets from fine mapping of the variants proximal to FOXA2 and the associated functional annotations.**

| rsID | MAF | p-value | OR (CI 95%) | Credible set | PIP | RegulomeDB score | RegulomeDB rank | CADD score |
| --- | --- | --- | --- | --- | --- | --- | --- | --- |
| rs1203884 | 0.14 | 2.19x10^-6^ | 0.64 (0.1-0.53) | Sentinel variant | - | 0.60906 | 4 | 17.39 |
| rs62205194 | 0.254334 | 0.883505 | 0.99 (0.85- 1.15) | 1 | 1 | 0.60906 | 4 | 0.702 |
| rs1999861 | 0.129531 | 0.740886 | 0.95 (0.68- 1.31) | 6 | 1 | 0.60906 | 4 | 0.095 |
| rs2021680 | 0.218046 | 0.00036218 | 0.66 (0.53- 0.83) | 2 | 1 | 0.55436 | 1f | 0.733 |
| rs1203867 | 0.121819 | 0.00010142 | 0.67 (0.54- 0.82) | 3 | 1 | 0.55436 | 1f | 1.957 |
| rs6137690 | 0.236884 | 0.691393 | 1.03 (0.89- 1.2) | 5 | 1 | 0.60906 | 4 | 9.485 |

**Supplementary Table 6. Significant eQTLs identified for credible set variants related to the PPFIA2 locus.**

| Gene | Variant | p-value | NES | Tissue |
| --- | --- | --- | --- | --- |
| PPFIA2 | rs2471500 | 1.70E-05 | -0.24 | Esophagus - Muscularis |
| PPFIA2 | rs2471500 | 9.00E-05 | -0.16 | Breast - Mammary Tissue |
| PPFIA2 | rs2471500 | 0.00011 | -0.18 | Adipose - Visceral (Omentum) |
| PPFIA2 | rs2471500 | 0.00035 | -0.14 | Nerve - Tibial |
| PPFIA2 | rs2471500 | 0.00053 | -0.16 | Cells - Cultured fibroblasts |
| PPFIA2 | rs2464738 | 2.20E-05 | -0.23 | Esophagus - Muscularis |
| PPFIA2 | rs2464738 | 0.00014 | -0.18 | Adipose - Visceral (Omentum) |
| PPFIA2 | rs2464738 | 0.00016 | -0.15 | Breast - Mammary Tissue |
| PPFIA2 | rs2464738 | 0.00033 | -0.14 | Nerve - Tibial |
| PPFIA2 | rs2464738 | 0.00053 | -0.16 | Cells - Cultured fibroblasts |
| PPFIA2 | rs7134579 | 1.80E-05 | -0.23 | Esophagus - Muscularis |
| PPFIA2 | rs7134579 | 7.00E-05 | -0.16 | Breast - Mammary Tissue |
| PPFIA2 | rs7134579 | 0.00026 | -0.14 | Nerve - Tibial |
| PPFIA2 | rs4842301 | 2.20E-05 | 0.23 | Esophagus - Muscularis |
| PPFIA2 | rs4842301 | 0.00014 | 0.18 | Adipose - Visceral (Omentum) |
| PPFIA2 | rs4842301 | 0.00016 | 0.15 | Breast - Mammary Tissue |
| PPFIA2 | rs4842301 | 0.00033 | 0.14 | Nerve - Tibial |
| PPFIA2 | rs4842301 | 0.00053 | 0.16 | Cells - Cultured fibroblasts |
| PPFIA2 | rs10083046 | 3.50E-05 | 0.23 | Esophagus - Muscularis |
| PPFIA2 | rs10083046 | 0.00015 | 0.18 | Adipose - Visceral (Omentum) |
| PPFIA2 | rs10083046 | 0.00038 | 0.14 | Nerve - Tibial |
| PPFIA2 | rs10083046 | 0.00055 | 0.16 | Cells - Cultured fibroblasts |

**Supplementary Table 7.** HaploReg annotations for credible set variants related to the PPFIA2 locus.

| rsID | Promoter histone marks | Enhancer histone marks | DNAse | Proteins bound | Regulatory motifs altered |
| --- | --- | --- | --- | --- | --- |
| rs2471500 | None | None | None | None | DMRT5, Irf_disc3, Irf_known3, Irf_known9, Pax-4_5, STAT_disc3 |
| rs2464738 | None | None | None | None | YY1_known2 |
| rs7134579 | None | None | None | None | CEBPG, Hoxa4, Irf_known7, Isx, Nkx2_8, Pax-8_1, Pax-8_2, Pou4f3, Sox_3 |
| rs4842301 | None | None | None | None | COMP1, EWSR1-FLI1 |
| rs10083046 | None | None | None | None | Gfi1_1, Spz1_1 |

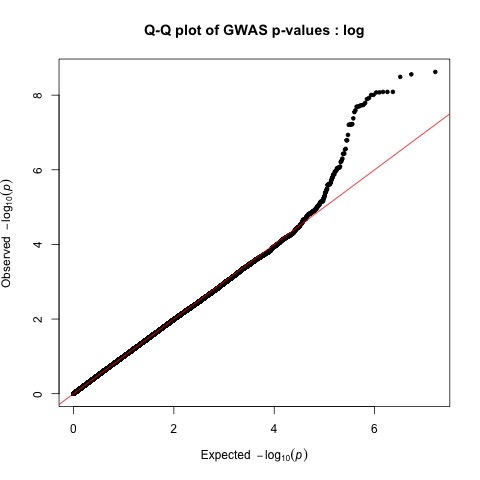

**Supplementary Figure 1**. Quantile-quantile (Q-Q) plot comparing the observed -log10 p-values from the GWAS on the y-axis with the expected p-values under the null hypothesis on the x-axis.

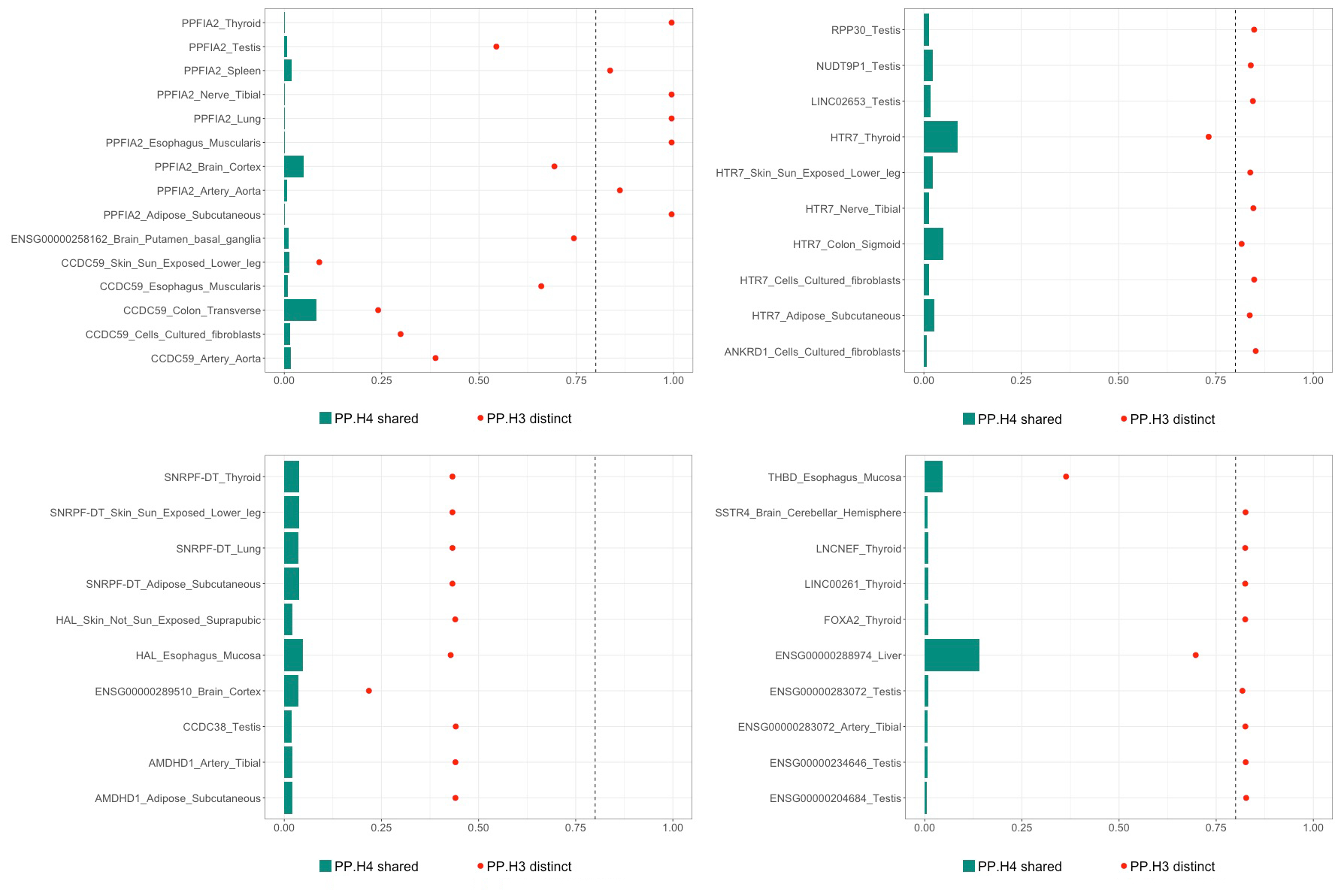

**Supplementary Figure 2**. Colocalisation results for rs9634233 and eQTLs from the GTEx v10 database.

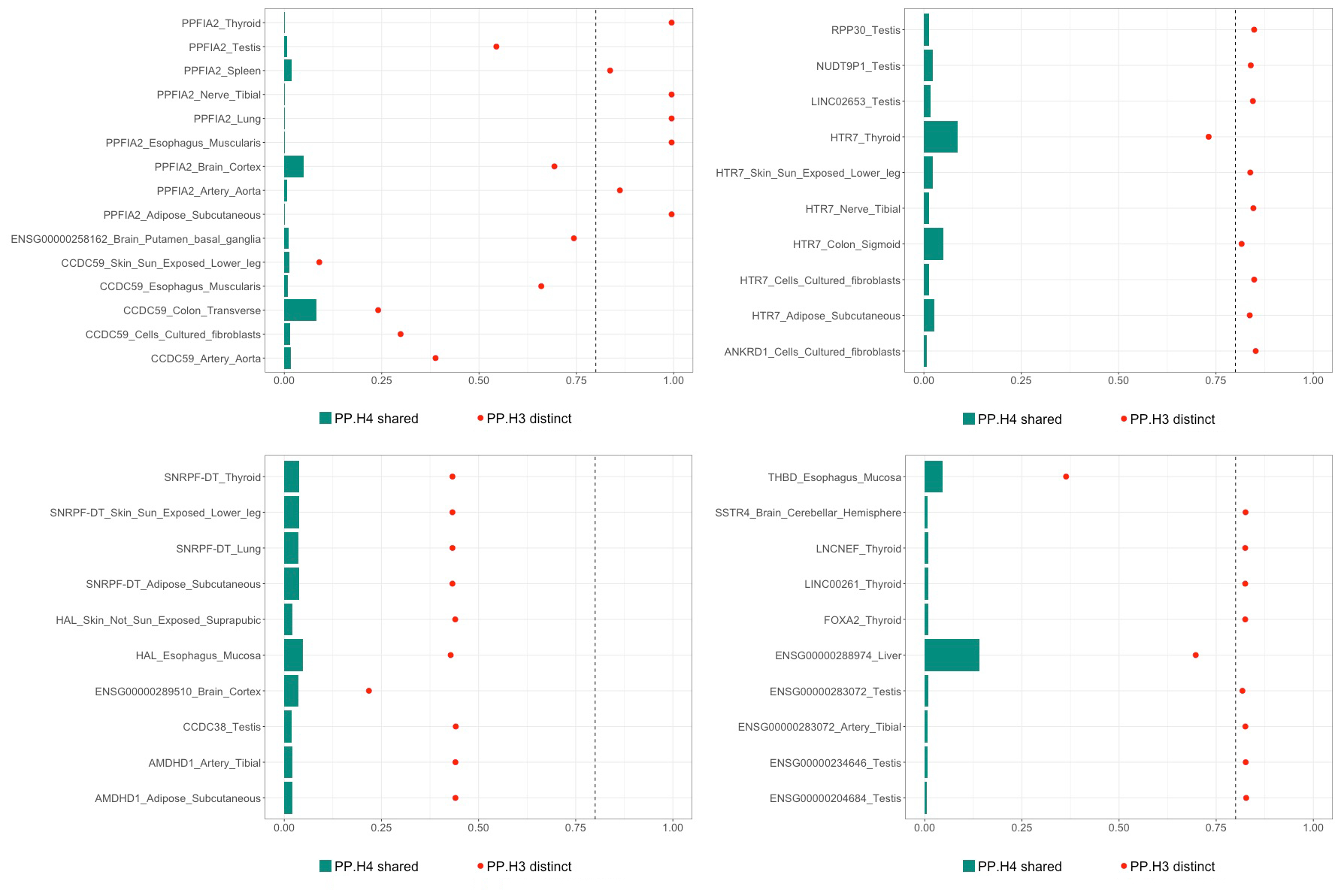

**Supplementary Figure 3.** Colocalisation results for rs17098642 and eQTLs from the GTEx v10 database.

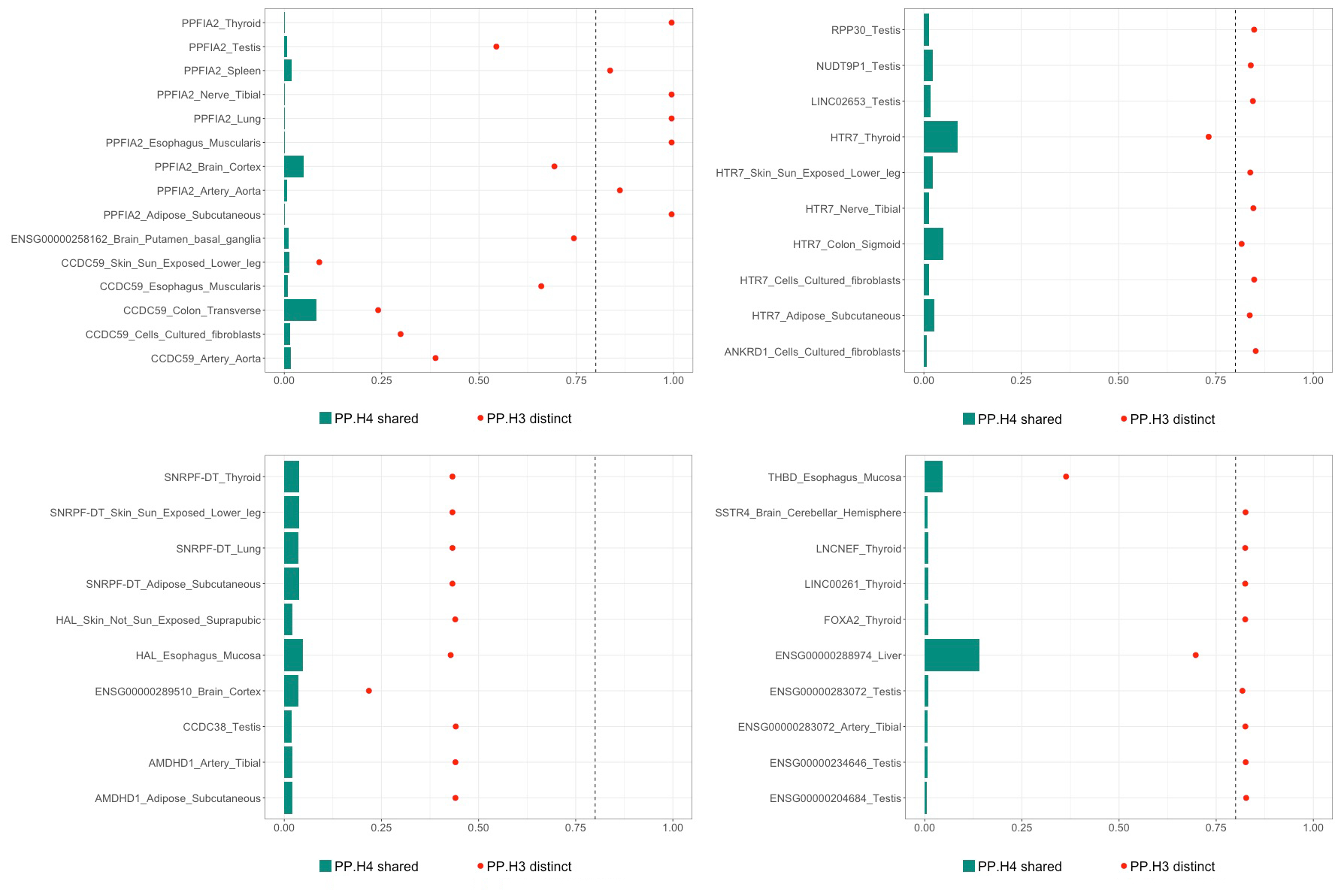

**Supplementary Figure 4.** Colocalisation results for rs75742107 and eQTLs from the GTEx v10 database.

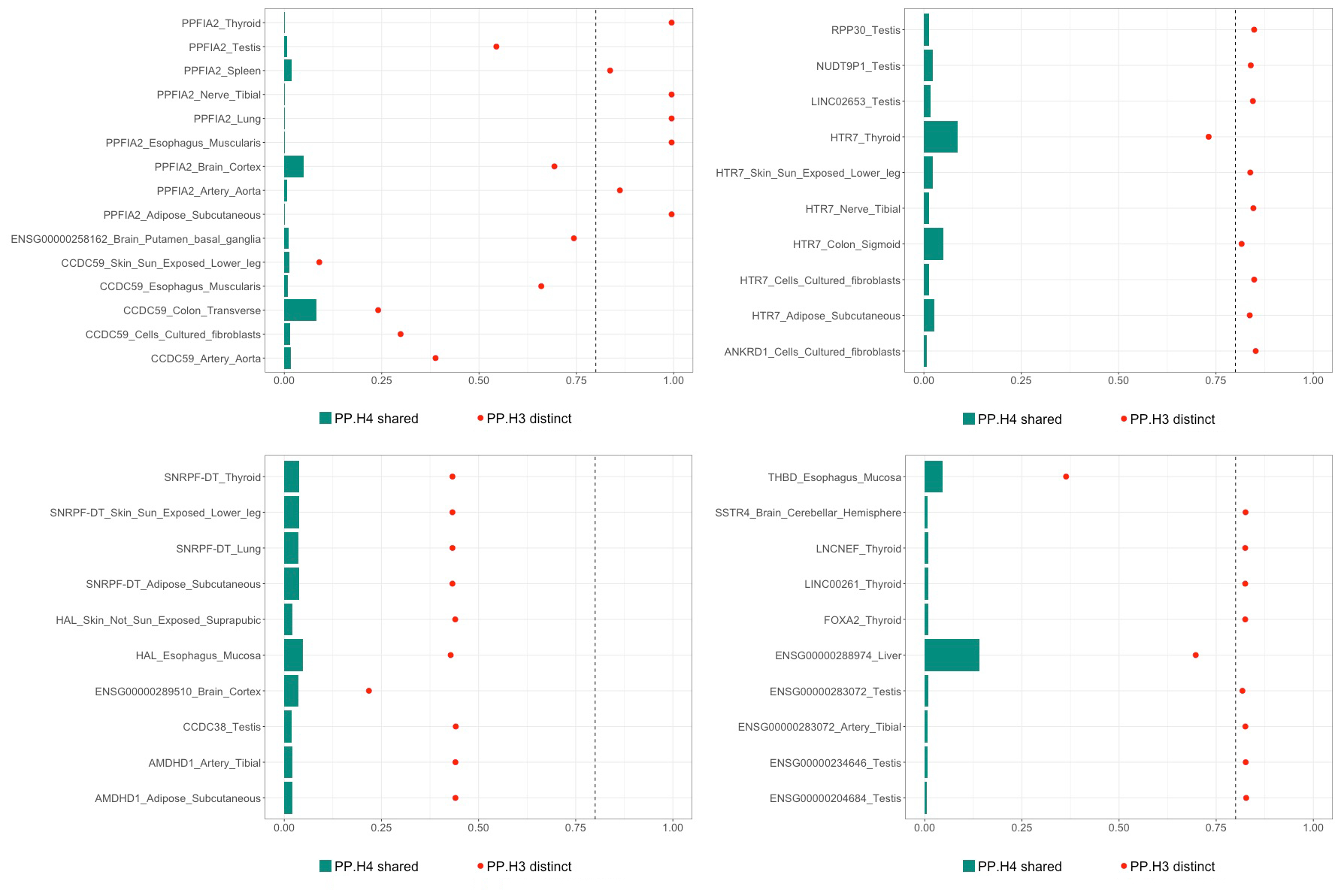

**Supplementary Figure 5.** Colocalisation results for rs1203884 and eQTLs from the GTEx v10 database.

**A**

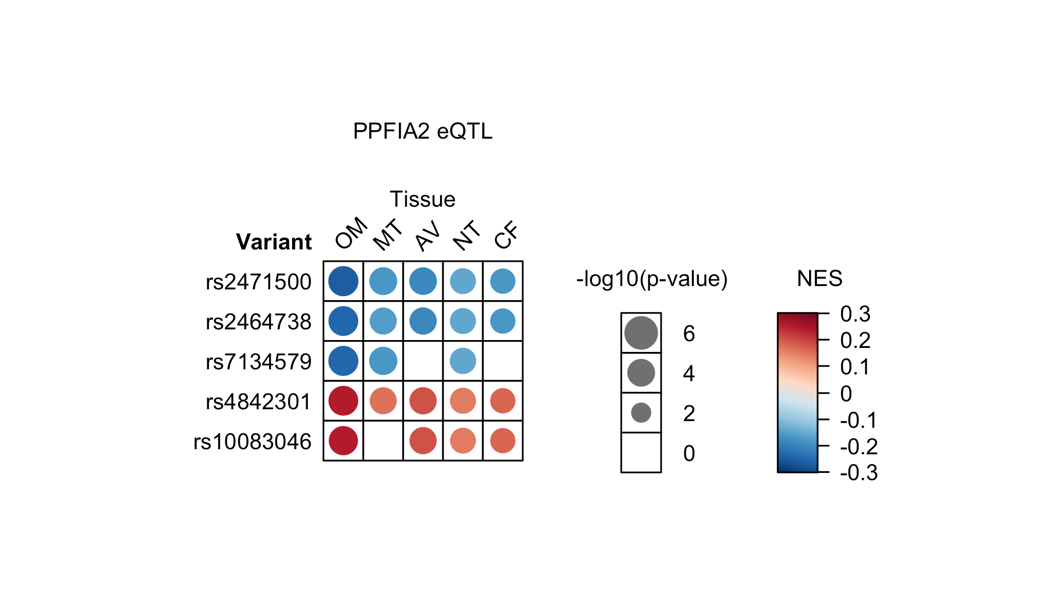

**B**

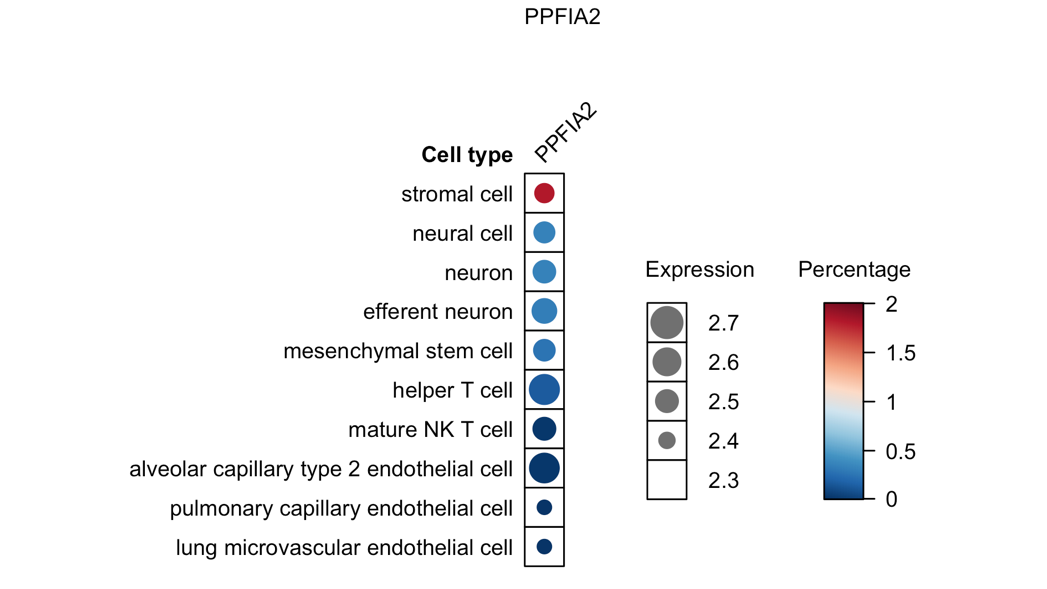

**Supplementary Figure 6.** Functional analyses of *PPFIA2* locus credible set variants using eQTL (A) and single cell gene expression of PPFIA2 in respiratory tract tissues (B). NES=normalised enrichment score. OM=oesophagus muscularis, MT=mammary tissue, AV=adipose visceral, NT=tibial nerve, and CF=cultured fibroblasts. NES=normalised enrichment score. Blank boxes= no significant eQTL identified

**A**

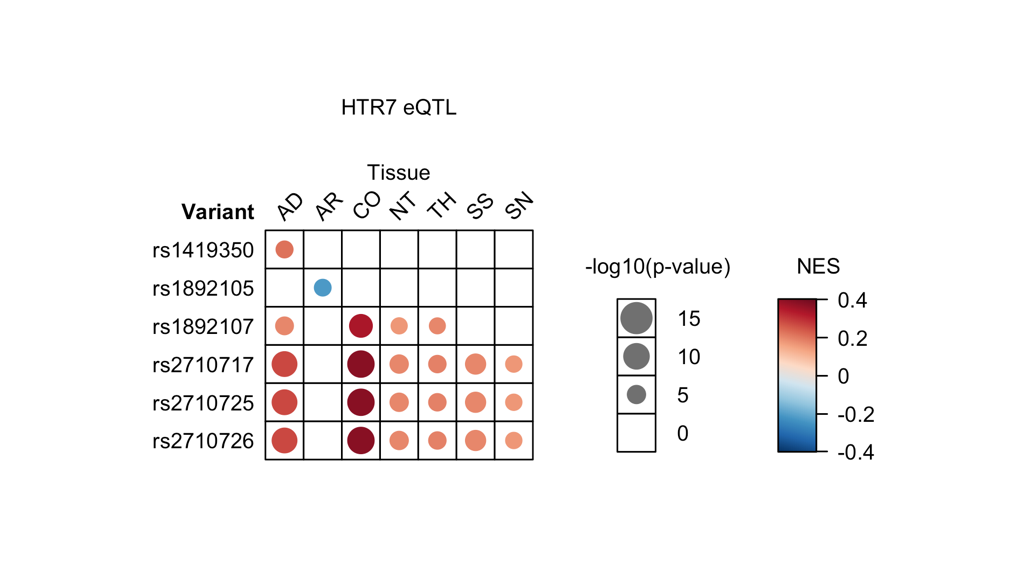

**B**

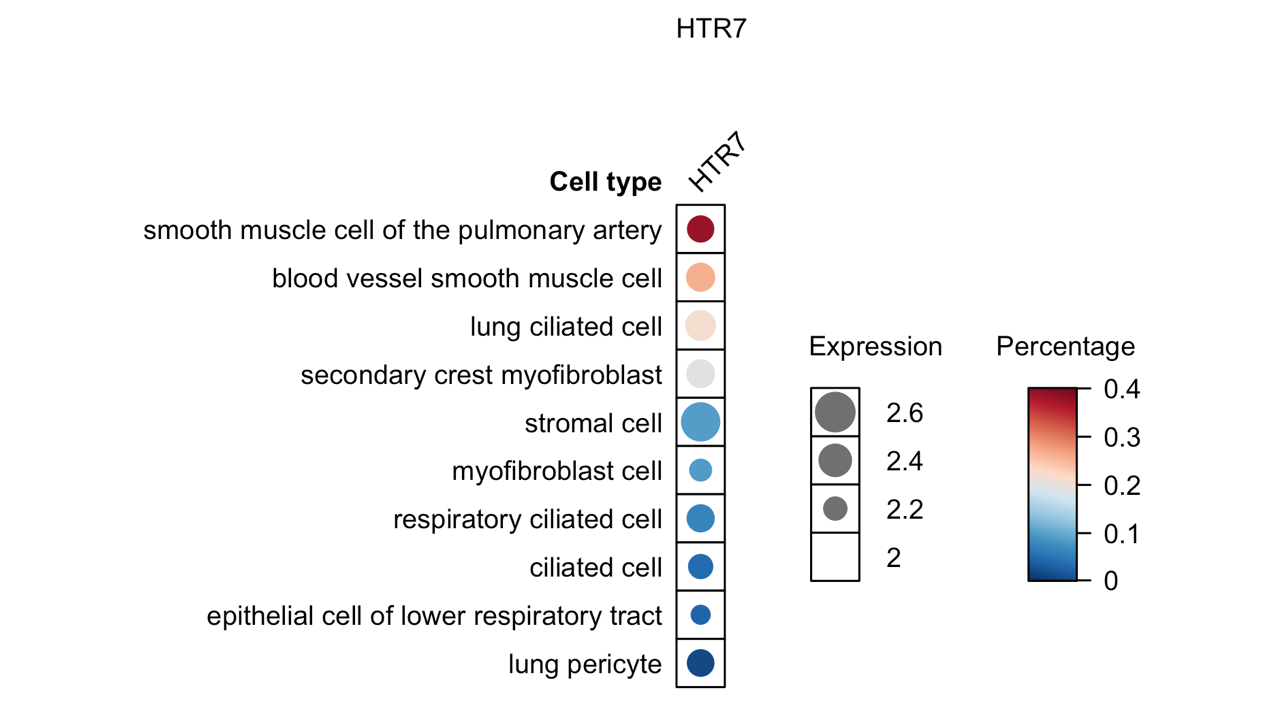

**Supplementary Figure 7.** Functional analyses of HTR7 locus credible set variants using eQTL (A) and single cell gene expression (B). AD=adipose (subcutaneous), AR=artery (tibial), CO= colon (sigmoid), NT=tibial nerve, TH=thyroid, SS=skin (sun exposed), and SN=skin (not sun exposed). NES=normalised enrichment score. Blank boxes= no significant eQTL identified.

**A**

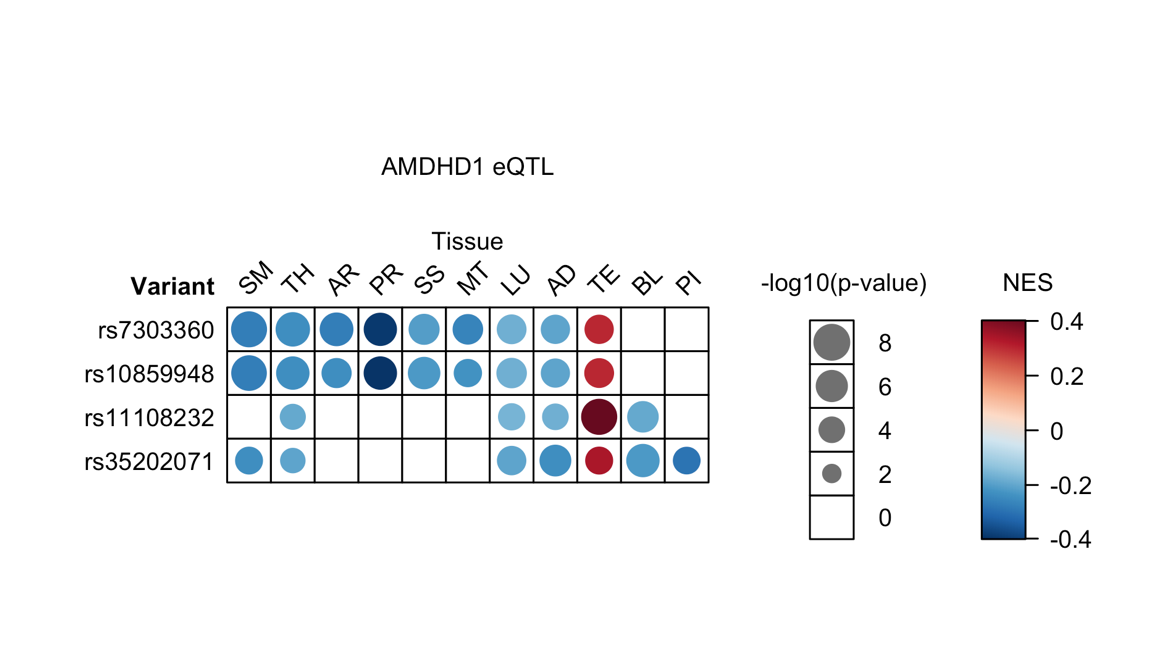

**B**

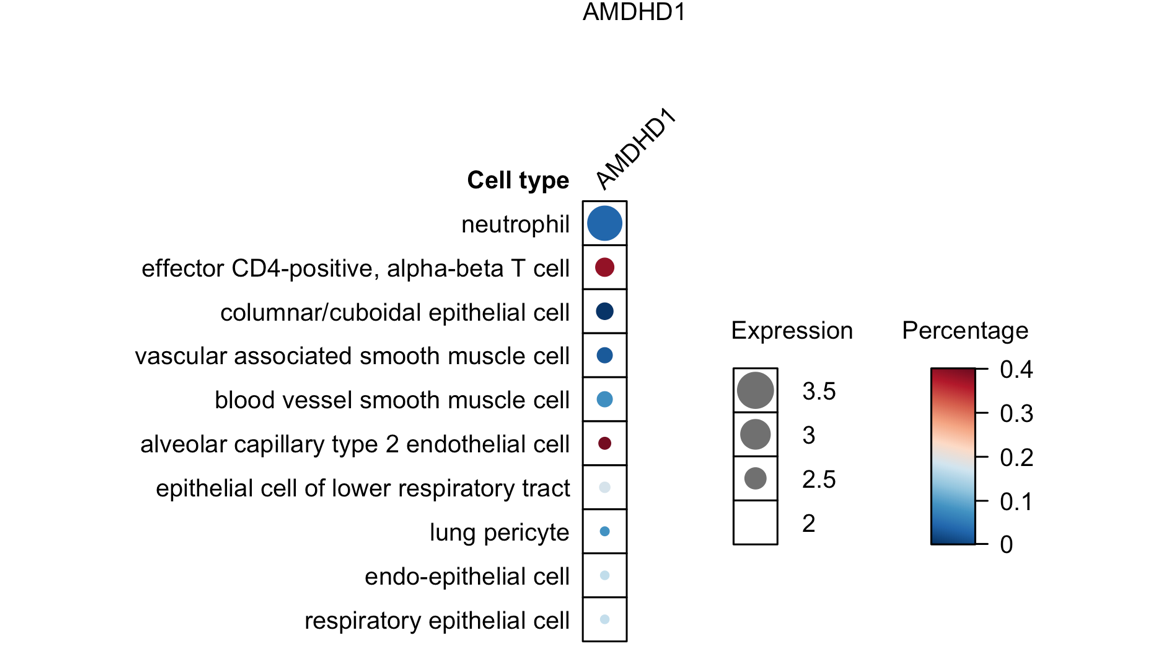

**Supplementary Figure 8.** Functional analyses of NTN4 locus credible set variants showing significant eQTLs to AMDHD1 (A) and single cell gene expression (B). AD=adipose (subcutaneous), AR=artery (tibial), CO= colon (sigmoid), NT=tibial nerve, TH=thyroid, SS=skin (sun exposed), and SN=skin (not sun exposed). NES=normalised enrichment score. Blank boxes= no significant eQTL identified.

**A**

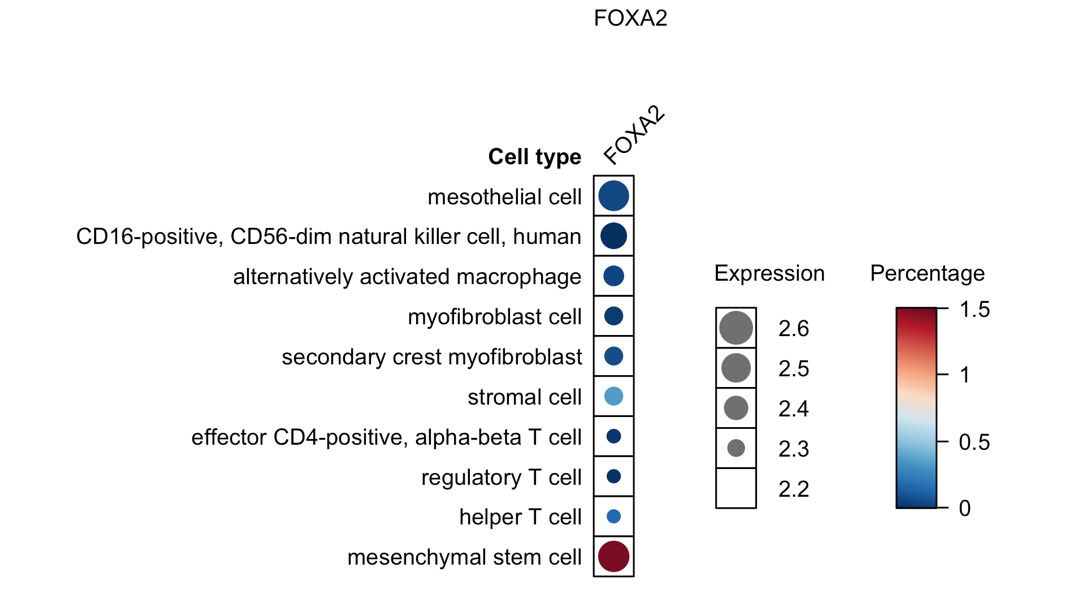

**Supplementary Figure 9.** Functional analyses of FOXA2 locus credible set variants showing single cell gene expression (A)
